# Immune Defense Protein IDP® Improves Cough and Chest Congestion During Cold and Flu Season in Older Adults: A Randomized, Double-Blind, Placebo-Controlled Study

**DOI:** 10.64898/2026.11.09.26367685

**Authors:** Azure D. Grant, Cypress S. Prideaux, Miguel Rosales, Lisa Capano-Wehrle, Jordan Moos, Nicole Mkrtytchian, Veronica Luna, Marie Crisel E. Krupa, Noah Craft

**Author notes:** **Corresponding Author:** Azure Grant, 11307 Hindry Ave, Suite D, Los Angeles, CA, 90034.

## Abstract

**Background:** Upper respiratory infections are common and carry a greater burden in older adults. IDP^®^ (Immune Defense Protein) is a dietary supplement whose active ingredient is a complex of milk-derived bioactive whey proteins. Prior studies of IDP^®^ report enhanced antibody response following influenza vaccination, suggesting a potential for reduction of incidence, duration, and severity of upper respiratory symptoms. This study evaluated the effects of IDP^®^ supplementation on the incidence, type, duration, and severity of upper respiratory symptoms in older adults during cold and flu season, along with rescue medication use and quality of life.

**Methods:** This was a randomized, double-blind, placebo-controlled study with three arms (placebo, standard dose of 50mg IDP® /day, and advanced dose of 200mg IDP® /day). Participants completed a one-week baseline period followed by a 16-week product use period, reporting daily upper respiratory symptoms via the Wisconsin Upper Respiratory Symptom Survey (WURSS-24). Participants additionally logged quality of life using the Short Form 36 (SF-36). ANCOVA for change from baseline, mixed models for repeated measures (MRMM), as well as Kruskal Wallis (KW) tests of area under the curve of symptom frequency and severity were used to compare trajectories over time.

**Results:** N=255 participants (n=88 Placebo, n=85 50 mg IDP®, n=82 200 mg IDP®) were evaluated. Analysis of individual symptom trends over time revealed that chest congestion and cough decreased in severity and duration significantly faster in the 50 mg IDP® and 200 mg IDP® groups compared to Placebo (KW for Area Under the Curve p<0.05). ANCOVA of baseline to Week-16 change and MRMM of total WURSS-24 symptom frequency, severity and rescue medication use revealed no significant difference between arms (ANCOVA and MRMM p > 0.05). SF-36 quality of life did not differ between arms. Adverse events potentially attributable to product use were infrequent, resolved, and did not differ between arms.

**Conclusions:** Sixteen weeks of daily IDP® supplementation was associated with a reduction in duration and severity of cough and chest congestion in older adults. IDP® appears safe and tolerable.

**Trial Registration:** The trial was reviewed and approved by BeyondBound IRB: BB2511NC-152, and registered at clinicaltrials.gov (NCT07431840)

## Introduction

Influenza and other upper respiratory infections are among the most common acute illnesses worldwide and a substantial cause of morbidity and mortality each year. Older adults are disproportionately affected (1), with heightened vulnerability due in part to immunosenescence, the age-related decline in innate and adaptive immune function that blunts the response to infection and vaccination (2,3).

Nutritional status is an important and modifiable determinant of immune function in older adults (4). In this population, nutritional supplementation both enhances antibody responses to influenza vaccination and reduces the number of days of upper respiratory symptoms (5,6), with specific benefits of prebiotic and milk-derived nutritional components (7,8).

IDP® (Immune Defense Protein) is a patented complex of over 50 milk-derived bioactive whey proteins, including lactoferrin and lactoperoxidase. These proteins have established antiviral activity against influenza and other respiratory and mucosal viruses (9,10). In animal models, IDP® enhances cell-mediated immune function, including delayed-type hypersensitivity and monocyte-macrophage phagocytic activity (11). In its first human trial, an 8-week double-blind, randomized, placebo-controlled pilot study, IDP® produced a dose-dependent enhancement of the antibody response to seasonal influenza vaccination in healthy adults. The high dose (200 mg/day) group exhibited relatively higher influenza antibody titer than placebo 2 weeks post-vaccination, and a significantly greater proportion of participants with a sustained four-fold antibody rise at 4 weeks (12).

However, enhanced vaccine immunogenicity does not necessarily translate into fewer or less severe respiratory symptoms in everyday life. The present randomized, double-blind, placebo-controlled study evaluated daily IDP® supplementation, at the same two doses studied previously (12). We hypothesized that IDP® consumption would be associated with fewer upper respiratory symptoms. Outcomes were the incidence, type, duration, and severity of upper respiratory symptoms (WURSS) in adults aged 60 and older across a full cold and flu season, together with evaluation of potential effects on rescue medication use, quality of life (SF-36), and tolerability.

## Methods

### Eligibility

Eligible participants were adults aged 60 years or older who experienced upper respiratory symptoms (i.e., coughing, sneezing, or congestion) at least two to three times during the cold and flu season and were able to read and understand English. Participants were required to provide informed consent, use a personal smartphone and download the Consumer Health Learning & Organizing Ecosystem (Chloe) by People Science app. They must have been able to receive product shipments within the United States and complete study assessments. Individuals were excluded if they lacked a personal smartphone or internet access, were unwilling to download Chloe, in an immunocompromised state including infectious, cancer-related, immunosuppressive-therapy, transplant, or autoimmune and inflammatory conditions, used investigational therapies within 30 days, had asthma or COPD, or had a diagnosed alcohol or substance use disorder. Participants were also ineligible if they had other underlying conditions or comorbidities that could confound the study outcomes, used cannabis- or nicotine-containing products daily, had known hypersensitivity to milk products, sorbitol, or tableting aids, or were judged by the principal investigator to be unable to comply with, or unsuitable for, the study.

### Study Design

This was a randomized, double-blind, placebo-controlled, three-arm study evaluating IDP® Preventative Supplementation against placebo in adults aged 60 and older during the cold and flu season. The study was conducted remotely through the Chloe by People Science app. Following screening and eligibility confirmation, participants were randomized in a blinded fashion to one of three arms (Placebo, 50 mg IDP, or 200 mg IDP) and were shipped study product within two weeks before the baseline period. During the one-week baseline period, participants completed a quality-of-life survey and were asked daily whether they had upper respiratory symptoms, completing the WURSS-24 along with rescue-medication and daily-activity questions when symptoms were present. During the 16-week product use period, participants took the assigned product or placebo once daily and completed daily well-being and symptom surveys, weekly adverse event questions, and monthly SF-36 assessments, followed by an end-of-study experience survey.

### Data Management System

All data was securely stored on Chloe Amazon Web Services HIPAA-compliant servers. The Chloe platform contains modules for building and managing surveys, study landing pages, marketing outreach with tracking tools for recruitment, audited electronic consent forms, data management and analytics using an integrated relational database. Additionally, the platform contains a user-facing app for participants that delivers surveys, study instructions, calendar reminders, communication with study team, and personal data reports at study culmination. Data from completed assessments was automatically collected for analysis.

### Recruitment, Consent, and Enrollment

Participants were recruited through social media channels and researcher networks. Recruitment outreach consisted of IRB-approved advertising by email, digital marketing channels and word of mouth. The IRB-approved study landing page hosted on the People Science website led to an IRB-approved pre-screening questionnaire to determine individual qualification. Qualified participants were invited to download the Chloe application (described below) and create an account to access the Informed Consent Form and continue through eligibility determination and enrollment.

Virtual electronic informed consent, including a study specific privacy authorization and the California Experimental Subject’s Bill of Rights (as applicable) were provided through the HIPAA-compliant cloud-based platform Chloe. Eligible participants who provided virtual electronic consent were automatically registered into the study by the platform.

Screening occurred within four weeks prior to enrollment; randomization and study product shipping occurred within two weeks prior to baseline. The daily survey covered general well-being and presence of upper respiratory symptoms; when symptoms were present, participants completed the WURSS-24 and additional questions on rescue-medication use and impact on daily activities. SF-36 was administered at baseline and monthly thereafter. Adverse events were assessed weekly.

### Study Timeline and Activities

Participants’ activities were as follows **(See Table 1 Study Timeline).**

**Table 1.**
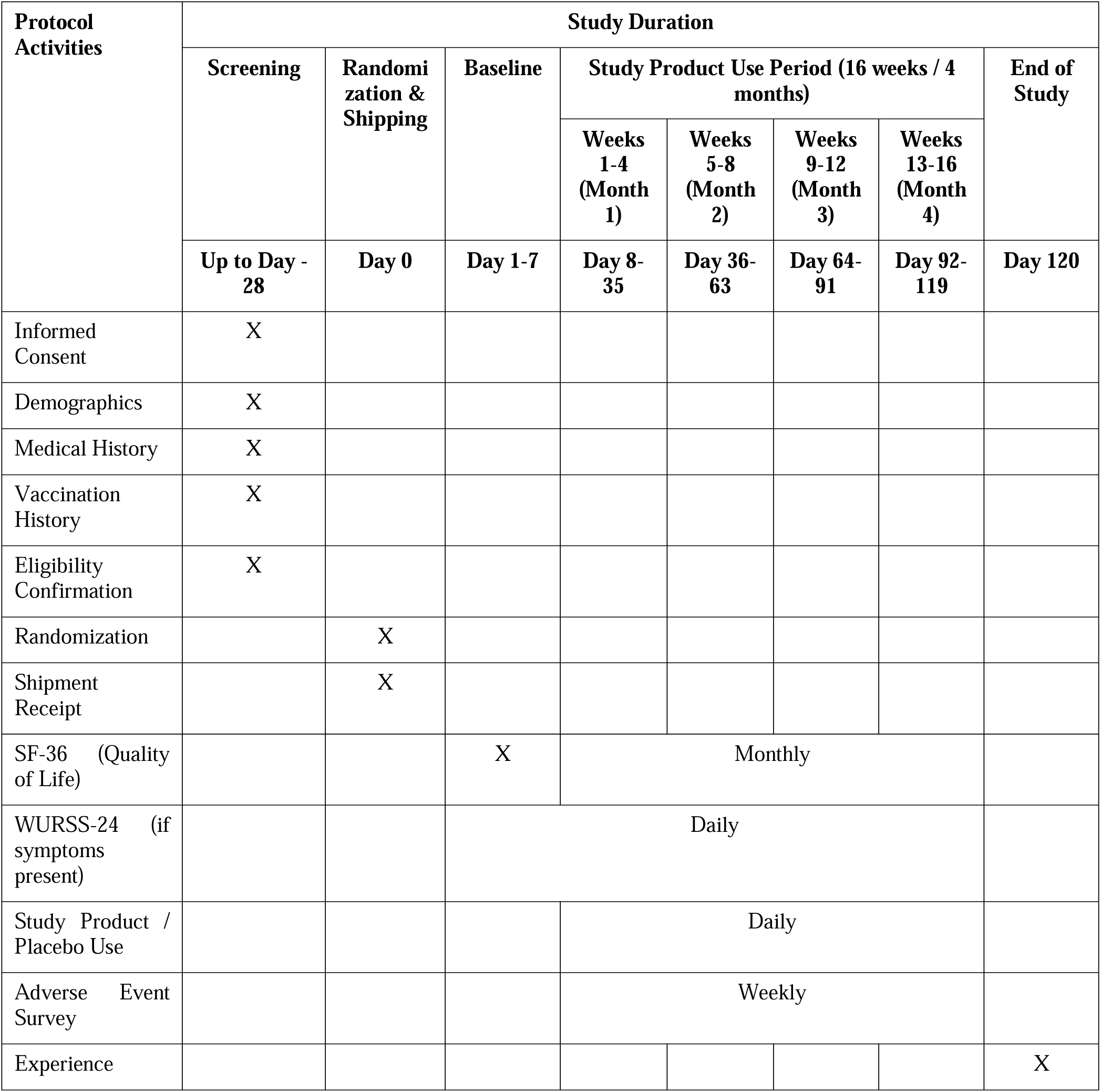
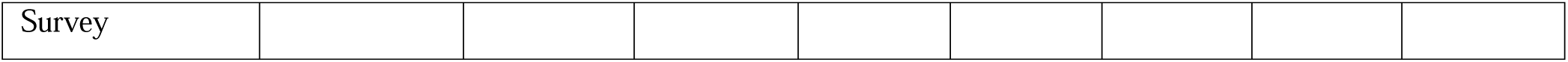
Study Activities.

| **Protocol Activities** | **Study Duration** |  |  |  |  |  |  |  |
|---|---|---|---|---|---|---|---|---|
|  | **Screening** | **Randomization & Shipping** | **Baseline** | **Study Product Use Period (16 weeks / 4 months)** |  |  |  | **End of Study** |
|  |  |  |  | **Weeks 1-4 (Month 1)** | **Weeks 5-8 (Month 2)** | **Weeks 9-12 (Month 3)** | **Weeks 13-16 (Month 4)** |  |
|  | **Up to Day -28** | **Day 0** | **Day 1-7** | **Day 8-35** | **Day 36-63** | **Day 64-91** | **Day 92-119** | **Day 120** |
| Informed Consent | X |  |  |  |  |  |  |  |
| Demographics | X |  |  |  |  |  |  |  |
| Medical History | X |  |  |  |  |  |  |  |
| Vaccination History | X |  |  |  |  |  |  |  |
| Eligibility Confirmation | X |  |  |  |  |  |  |  |
| Randomization |  | X |  |  |  |  |  |  |
| Shipment Receipt |  | X |  |  |  |  |  |  |
| SF-36 (Quality of Life) |  |  | X | Monthly |  |  |  |  |
| WURSS-24 (if symptoms present) |  |  | Daily |  |  |  |  |  |
| Study Product / Placebo Use |  |  |  | Daily |  |  |  |  |
| Adverse Event Survey |  |  |  | Weekly |  |  |  |  |
| Experience Survey |  |  |  |  |  |  |  | X |

### Intervention Preparation and Dosing

The 1g rectangular, rounded end tablets were produced by an approved NZ based health supplement manufacturer with the placebo formulation comprising Sorbitol, Microcrystalline cellulose, Chocolate powder, Milk Flavouring and Magnesium stearate. To produce the IDP® 50mg standard dose tablet, 50mg of microcrystalline cellulose was omitted from the placebo formula. To produce the IDP® 200mg advanced dose tablet, 160mg of microcrystalline cellulose and 40mg of chocolate powder were omitted. This resulted in visually similar tablets to support the blinding of investigators and patients. Dosing was 1 tablet per day in the morning with or without food.

### Subjective Daily Assessments

Each day, participants were asked whether they had any upper respiratory symptoms and, if so, completed the Wisconsin Upper Respiratory Symptom Survey (WURSS-24), rating symptom severity and functional impact. Participants also reported daily adherence to product use, any changes to daily medications, and consumption of alcohol or evening caffeine.

### Outcome Measures

The Wisconsin Upper Respiratory Symptom Survey (WURSS-24) was the primary outcome measure. The WURSS-24 is a validated 24-item instrument for assessing upper respiratory illness. It comprises a global severity item, a set of symptom-severity items (such as runny nose, plugged nose, sneezing, sore throat, scratchy throat, cough, hoarseness, head congestion, and chest congestion), and items assessing the impact of illness on daily function and quality of life, each rated on a 0 to 7 scale (0 = not sick or not at all affected, 7 = severe), with higher scores indicating greater symptom burden. Each day during the baseline period and the 16-week product use period, participants first reported whether they had any upper respiratory symptoms and, if so, completed the WURSS-24 along with questions on rescue medication use and impact on daily activities. Individual symptoms as well as a daily total symptom-burden score were computed as the sum of the scored symptom and functional items. From these daily reports, the frequency (proportion of days with any upper respiratory symptom), type (per-symptom prevalence), duration (length of each symptom episode), and severity (WURSS-24 burden) of upper respiratory symptoms were derived as the primary endpoints.

Secondary outcomes included Quality of Life (SF-36), rescue medication use, and an experience survey. Health-related quality of life was measured with the 36-Item Short Form Health Survey (SF-36), administered at baseline and monthly during the product use period, which yields eight subscale scores and an overall score, with higher scores indicating better quality of life. Rescue medication use, including the type and frequency of medications taken for upper respiratory symptoms, was recorded daily throughout the study.

### Adverse Events

Participants were prompted weekly to disclose any adverse events that occurred.

### Data Evaluability, Analysis and Statistics

Participants with at least three days of data in their baseline week and at least three days in the final product week (Week 16) were evaluated for baseline to end of study symptom analysis. WURSS-24 daily totals were recomputed from the raw items. Weekly symptom burden was defined per participant as the mean daily WURSS total, scored zero on symptom-free days. The primary between-arm comparison for severity was an ANCOVA of Week-16 burden adjusted for baseline symptom burden, age, and sex, supported by a mixed model for repeated measures across weeks. Between-arm change on frequency, duration, and severity was summarized with age- and sex-adjusted mixed models and tested with a robust Kruskal-Wallis test. Area under the curve was computed using the trapezoidal method and differences computed using ranksum tests (KW for all 3 groups, or Mann-Whitney U for combined 50 mg IDP® and 200 mg IDP® groups). Benjamini-Hochberg correction for multiple comparisons was applied across endpoints. Fisher’s Exact Test (FET) was used to compare percentages.

## Results

### Ethics Approval and Consent to Participate

This study was reviewed and approved by BeyondBound IRB BB2511NC-152, and all participants gave informed consent.

### Consent for Publication

No individual n=1 data, images or videos are reported in this manucript.

### Recruitment and Conduct

Participants were recruited from December 1st, 2025-March 1st, 2026, through social media and researcher networks and completed all study activities remotely using the Chloe app. Randomization was blindly and automatically 1:1:1 using the Chloe software. Each arm was assigned an equal target allocation proportion. At each allocation, Chloe compared the cumulative proportion of participants assigned to each arm against its target and assigned the participant to the first arm that was below its target. Across comparable three-arm studies conducted on the platform, allocation remained within three participants and two percentage points of the target proportions. Participants were enrolled formally by blinded study coordinators following randomization.

### Sample Size, Compliance and Demographics

Of 281 participants who were randomized, 265 completed the study and 16 were withdrawn due to loss to follow-up (7), participant decision (7), and investigator or other decision (2) (**Figure 1**). Reported daily product use adherence averaged 84.0%. N=255 participants were evaluable based on sufficient baseline and end of study data and analyzed. Groups did not differ significantly at baseline on age, sex, or race/ethnicity (all Fisher’s Exact Test p>0.05). Demographics are described in **Table 2**. Participants collected product use from December 2025 to June 2026, with the majority of data collected from February to April 2026. The trial was concluded when all randomized, non-withdrawn participants completed the study.

**Figure 1.**
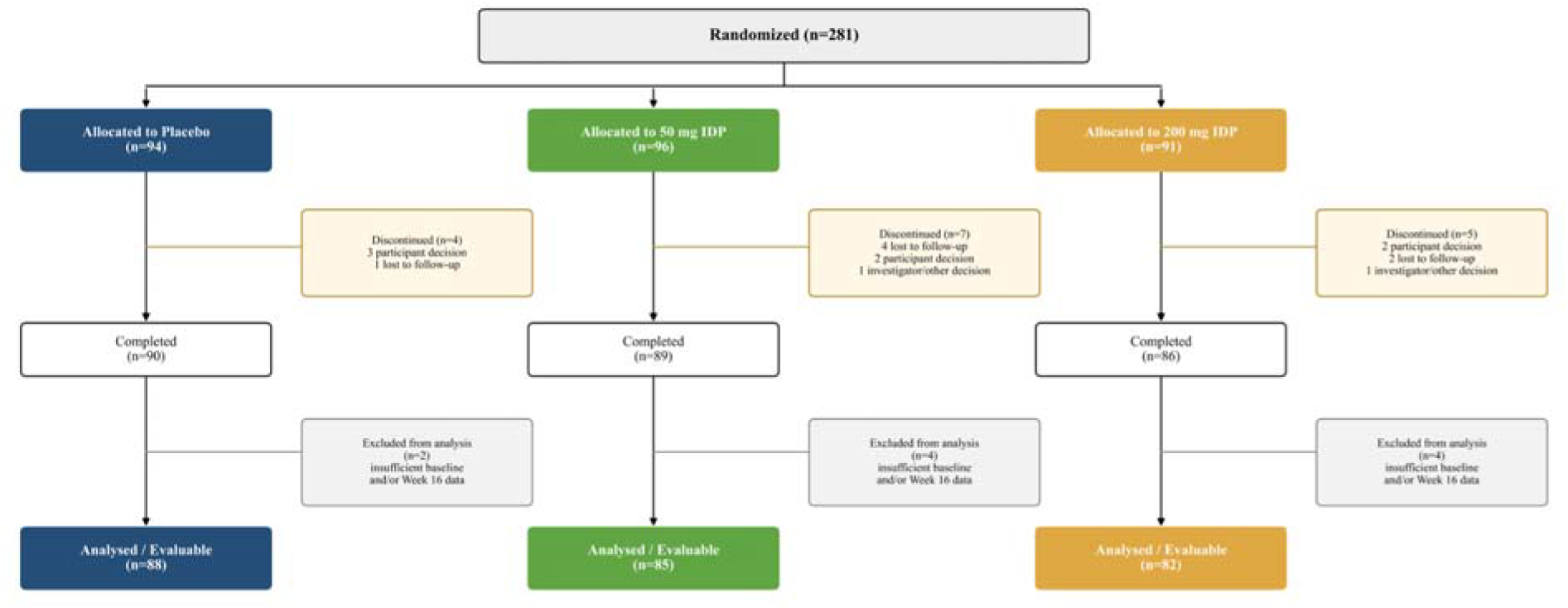
CONSORT Diagram.

**Table 2.**
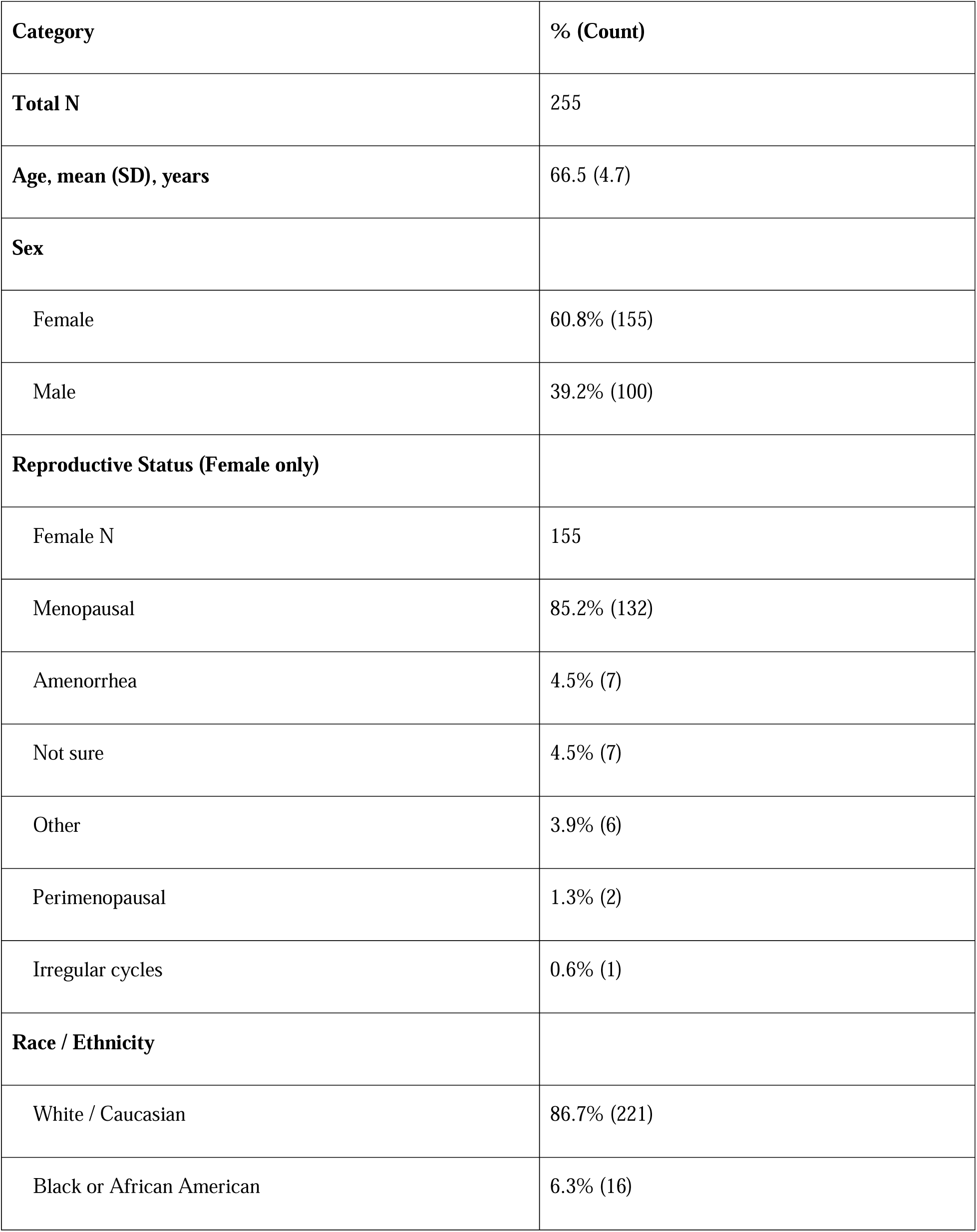
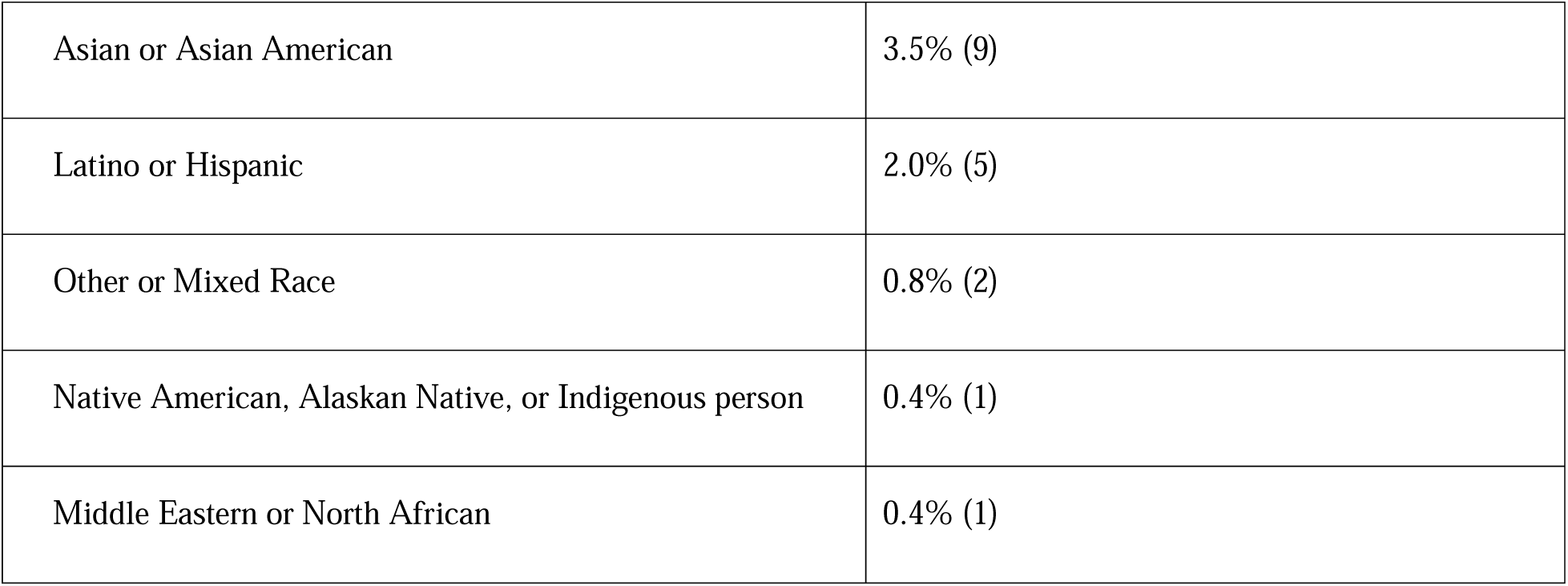
Demographics.

| **Category** | **% (Count)** |
|---|---|
| **Total N** | 255 |
| **Age, mean (SD), years** | 66.5 (4.7) |
| **Sex** |  |
| Female | 60.8% (155) |
| Male | 39.2% (100) |
| **Reproductive Status (Female only)** |  |
| Female N | 155 |
| Menopausal | 85.2% (132) |
| Amenorrhea | 4.5% (7) |
| Not sure | 4.5% (7) |
| Other | 3.9% (6) |
| Perimenopausal | 1.3% (2) |
| Irregular cycles | 0.6% (1) |
| **Race / Ethnicity** |  |
| White / Caucasian | 86.7% (221) |
| Black or African American | 6.3% (16) |
| Asian or Asian American | 3.5% (9) |
| Latino or Hispanic | 2.0% (5) |
| Other or Mixed Race | 0.8% (2) |
| Native American, Alaskan Native, or Indigenous person | 0.4% (1) |
| Middle Eastern or North African | 0.4% (1) |

### Upper Respiratory Symptom Incidence

Groups did not differ at baseline on symptom frequency or severity (both ANCOVA change from baseline and MMRM p>0.05), though a nominal baseline difference in symptom duration was observed (p=0.037). Symptom measures declined across the whole cohort over the product use period, and the decline was similar across arms. Between-arm change did not differ for frequency, duration, or severity (all ANCOVA change from baseline and MMRM p>0.05) (**Figure 2**). However, area under the curve of cough and chest congestion prevalence and severity were reduced at a significantly greater rate in the pooled 50 mg IDP® + 200 mg IDP® group (MWU BH-FDR p<0.05 for both measures, weeks 1-8 of product use, respectively) (**Figure 3**). IDP® doses did not differ from one another (MWU p>0.05). In the initial 4 weeks, this corresponded to a 51.4% drop in cough and a 53.3% drop in chest congestion. Over 8 weeks, this corresponded to a total 34.2% drop in cough AUC and a 37.3% drop in chest congestion. There was no difference between the 50mg and 200 mg doses for either symptom (cough MWU p=0.97 and chest congestion MWU p=0.67).

**Figure 2.**
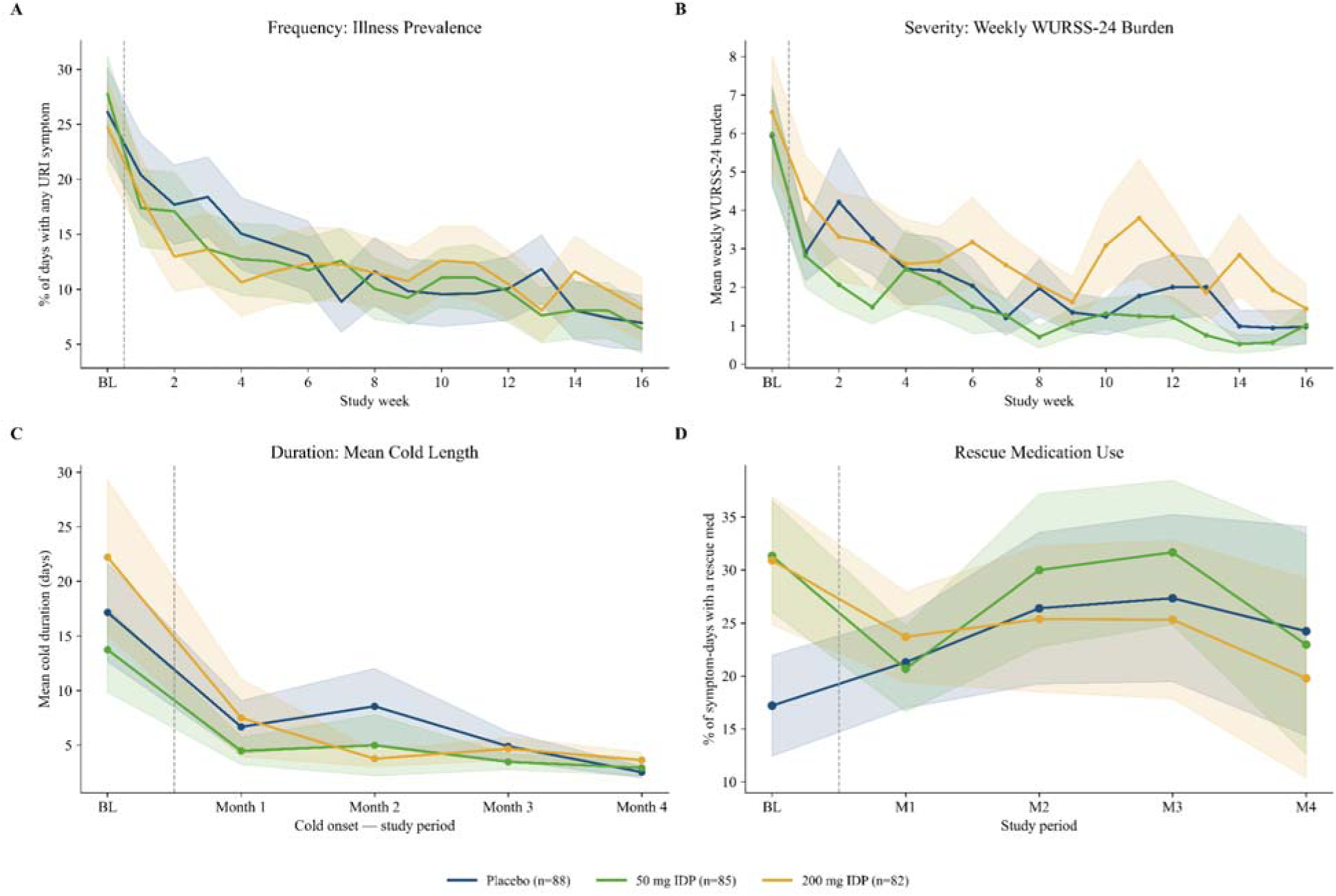
Total URI Symptom Burden Frequency, Severity and Duration Improve Comparably Across Groups, While Rescue Medication Use Frequency Does Not Change. Mean with shaded SEM A) Symptom Frequency, B) Symptom Severity, C) Symptom Duration and D) Rescue Medication Use across the study period. Placebo is shown in blue, 50 mg IDP® in Green and 200 mg IDP® in Orange. ANCOVA p>0.05 and MMRM p>0.05 for all metrics.

**Figure 3.**
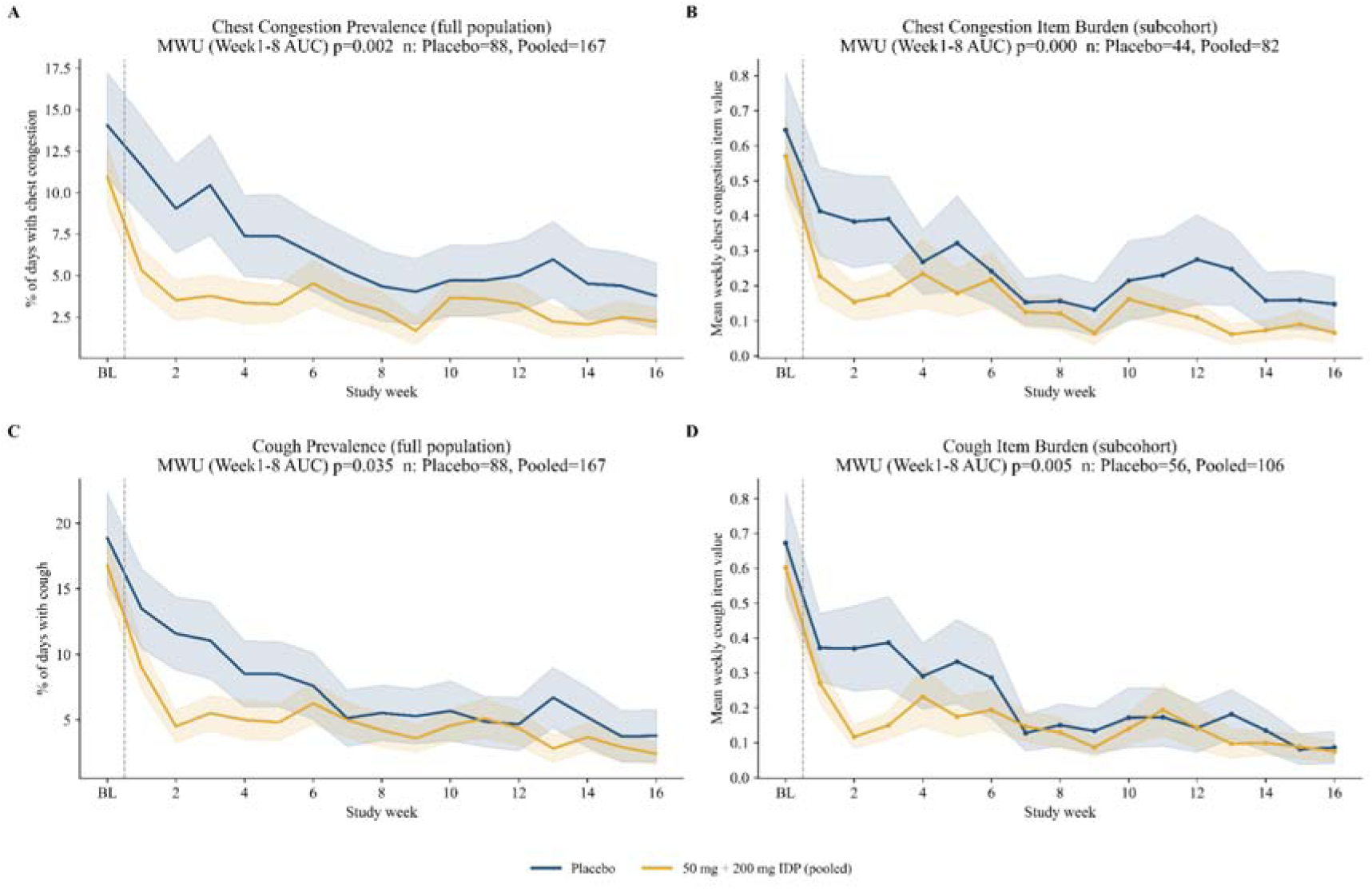
Pooled Analysis of the 50 mg IDP® and 200 mg IDP® groups Suggests Faster Improvement in Symptom Type: Per-Symptom Prevalence Trajectories. Mean with shaded SEM of trajectories of A) Chest congestion prevalence, B) Chest congestion severity, C) Cough prevalence and D) Cough severity across the study period in Placebo (blue) vs. pooled 50 mg IDP® and 200 mg IDP® (orange). Cough and chest congestion reduce at a significantly greater rate in the pooled 50 mg IDP® + 200 mg IDP® group in prevalence and severity. Pooled prevalence indicates prevalence % of all logged symptoms per participant. Item burden or severity is evaluated among participants who reported the presence of that symptom. MWU indicates that the pooled 50 mg IDP® + 200 mg IDP® group has significantly lower area AUC for chest congestion and cough prevalence and severity (MWU FDR-BH p<0.05 for all).

Data on symptom trajectories were additionally segregated by whether participants were asymptomatic at baseline, but no statistical group differences were noted (data not shown). No group differences emerged by demographic factors (sex, age, ethnicity, data not shown).

### Quality of Life (SF-36)

The 50 mg IDP® and 200 mg IDP® groups exhibited a small trend toward increased quality of life total score +3.4 SD 9.4 points in B, +3.1 SD 12.2 points in C (ANCOVA p=0.049). Groups did not differ significantly at any time point (MMRM p>0.05). No demographic factors impacted these results **(See Figure 3)**.

**Figure 4.**
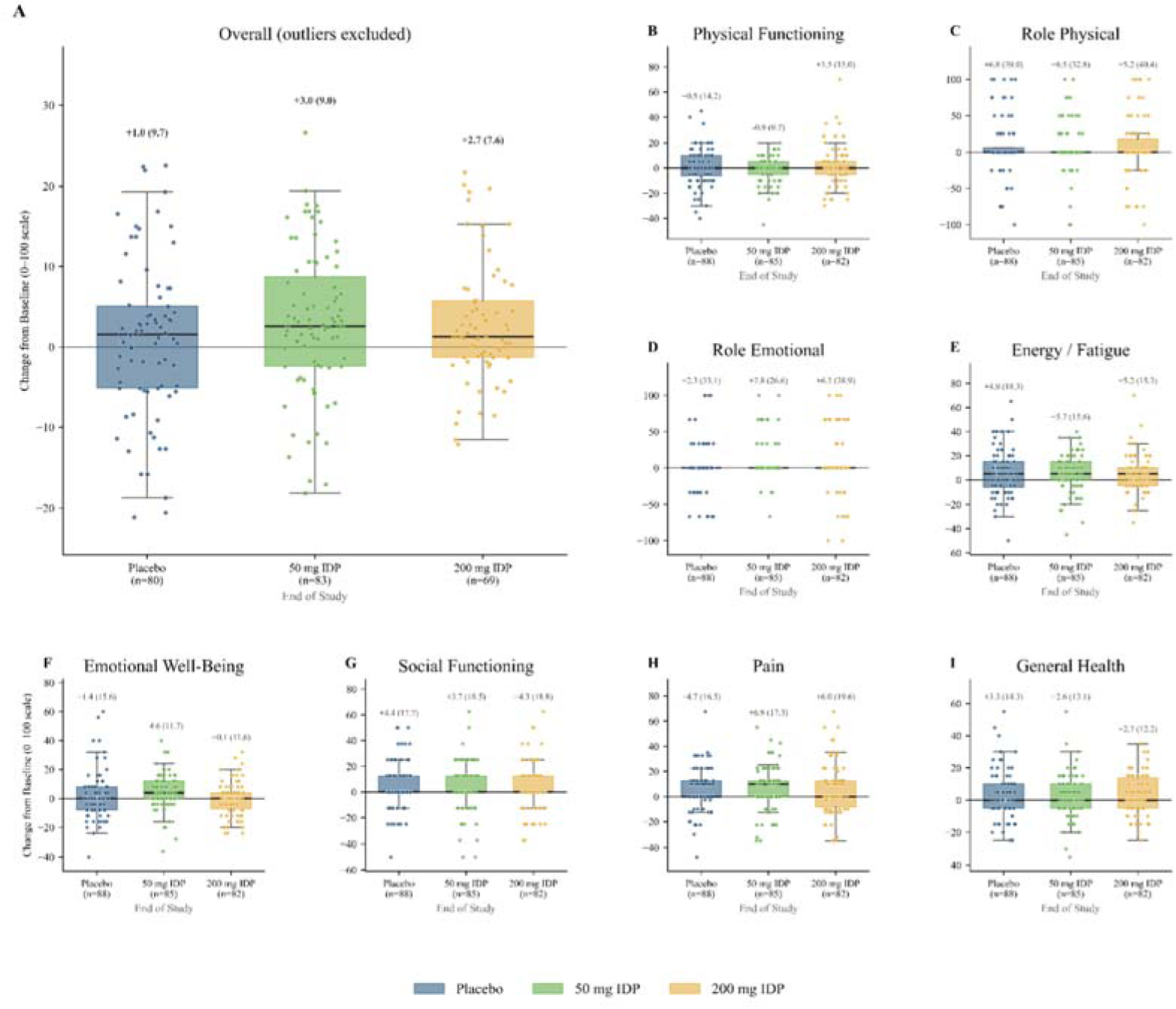
Change in SF-36 Quality of Life Trends Toward Improvement in the 50 mg IDP® and 200 mg IDP® groups. Box and whiskers with overlaid scatters of individual data of Placebo (blue), 50 mg IDP® (green) and 200 mg IDP® (yellow), reveal a small, non-statistical trend toward greater improvement in the 50 mg IDP® and 200 mg IDP® groups to end of study for overall sore (A). Subscores are shown in (B-I).

#### Adverse Events

For adverse events unlikely to be related to study product use, no AE differed in prevalence by group. Overall, 43 of the randomized participants (15.3%) reported at least one adverse event (68 events total), with no significant between-arm difference in the proportion reporting an AE (p > 0.05). 3 serious adverse events were reported but not considered possibly related to study product use: a fall down the stairs and an Achilles tendon tear at different time in the same individual (200 mg IDP), and an episode of vertigo determined to be caused by displaced otoliths (50 mg IDP). Of note, an episode of spleen or rib cage pain was reported as severe by the participant and resulted in withdrawal. The AE was marked as potentially related to product use as the participant reported that the pain was resolved after product use cessation. There were no follow up episodes of pain. Severity and potential relationship to product are summarized in Table 4.

**Table 4.** Adverse Events Potentially Related to Product Use: Safety Summary by Type.

| AE Name | Group | Count | Unique Participants | % Mild | % Moderate | % Severe |
|---|---|---|---|---|---|---|
| Itching Mouth | 200 mg IDP | 2 | 1 | 100.0 | 0.0 | 0.0 |
| Diarrhea | 200 mg IDP | 1 | 1 | 0.0 | 100.0 | 0.0 |
| Headache | 50 mg IDP | 1 | 1 | 0.0 | 100.0 | 0.0 |
| Spleen Pain/Left Rib Cage Pain | 50 mg IDP | 1 | 1 | 0.0 | 0.0 | 100.0 |

## Discussion

Sixteen weeks of daily IDP® supplementation in adults aged 60 and older was associated with significantly lower prevalence of cough and chest congestion relative to placebo. Interestingly, the doses performed comparably, suggesting 50 mg is sufficient to produce this effect. Upper respiratory symptom burden declined markedly from baseline through the product-use period across the cohort as a whole, as expected for participants enrolled during peak symptom season and followed as the season recedes, and potentially further amplified by regression to the mean among individuals who joined the study while symptomatic. Quality of life improved modestly more in both IDP® arms than placebo, though the difference did not reach statistical significance. IDP® was well-tolerated: adverse events were infrequent, did not differ by arm, and the one event considered possibly product-related was resolved after discontinuation.

These findings are consistent with the immunologic rationale for IDP. The preceding trial demonstrated that IDP® enhances the antibody response to influenza vaccination in a dose-related manner (12). Cough and chest congestion are the symptoms most directly tied to lower-respiratory viral burden, where an antibody-mediated effect would be expected to matter most. In addition, experimental studies of milk-derived proteins have described immunomodulatory and antiviral activities, while a mouse influenza model found that orally administered lactoferrin or lactoperoxidase reduced lung consolidation and inflammatory-cell infiltration (13).

This study has several limitations. These include a predominantly Caucasian study population, which may limit generalizability. Additionally, this study was a decentralized clinical trial, intentionally designed to collect data at home during the everyday lives of participants. Participants’ measurements were obtained using an app-based interface. This design was chosen to evaluate the impacts of IDP® under real-world conditions with minimal compromise on data quality. Due to the nature of real-world conditions, this dataset may exhibit greater variability due to within-individual differences and variable outside circumstances in comparison to traditional site-based clinical trials (14). Finally, all symptoms were self-reported, and antibody data were not collected.

## Conclusions

Daily IDP® supplementation over sixteen weeks was safe, well-tolerated and associated with faster improvement in cough and chest congestion in older adults. Future trials are needed to formally evaluate equivalency among 50mg and 200mg doses, as well as impact on objective immune markers in naturally acquired illness in a real-world setting.

## Author Contributions

Conceptualization: AG, MCEK, NC. Data curation: ADG, CP, PLO. Formal analysis & visualization: ADG, CSP. Funding acquisition: NC, MCEK. Investigation: MCEK, ADG, VL, JM, NM, MR, LCW, NC. Manuscript preparation: ADG, CP, NC. All authors read and approved the final manuscript.

## Acknowledgements

N/A

## Data Availability

Data are available upon reasonable request from the corresponding author.

## Abbreviation

AE: Adverse Event
ANCOVA: Analysis of Covariance
App: Application
AUC: Area Under the Curve
CFR: Code of Federal Regulations
Chloe: Consumer Health Learning and Organizing Ecosystem
CI: Confidence Interval
COPD: Chronic Obstructive Pulmonary Disease
FDR: False Discovery Rate
GCP: Good Clinical Practice
HIPAA: Health Insurance Portability and Accountability Act
ICH: International Council for Harmonisation
IDP®: Immune Defense Proteins
IRB: Institutional Review Board
MMRM: Mixed Model for Repeated Measures
NIH: National Institutes of Health
OTC: Over the Counter
PHI: Protected Health Information
PI: Principal Investigator
SAE: Serious Adverse Event
SD: Standard Deviation
SEM: Standard Error of the Mean
SF-36: 36-Item Short Form Health Survey
URI: Upper Respiratory Infection
VAS: Visual Analogue Scale
WURSS-24: Wisconsin Upper Respiratory Symptom Survey, 24-item

## Supplemental Materials

**Supplemental Figure 1:**
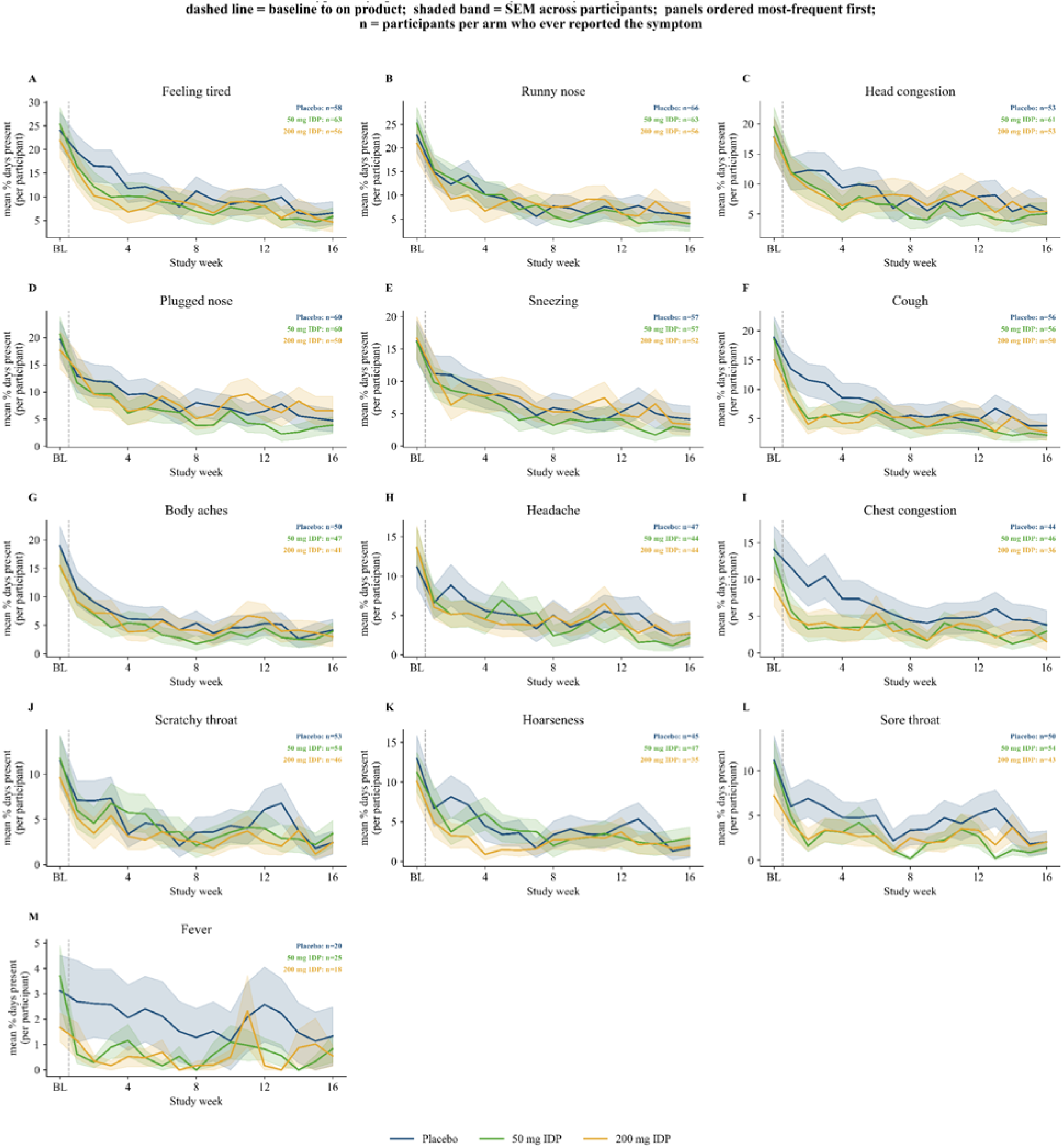
Symptom Prevalence Trajectory by Group Suggested More Rapid Symptom Abatement in Cough and Chest Congestion. Mean with SEM shaded by group of symptom trajectories across the study. Placebo is represented in blue, 50 mg IDP® in green, 200 mg IDP® in yellow. Symptoms are ordered from most to least prevalent as follows: A) Feeling Tired, B) Runny nose, C) Head congestion, D) Plugged nose, E) Sneezing, F) Cough, G) Body aches, H) Headache, I) Chest congestion, J) Scratchy throat, K) Hoarseness, L) Sore throat, M) Fever.

**Supplemental Figure 2.**
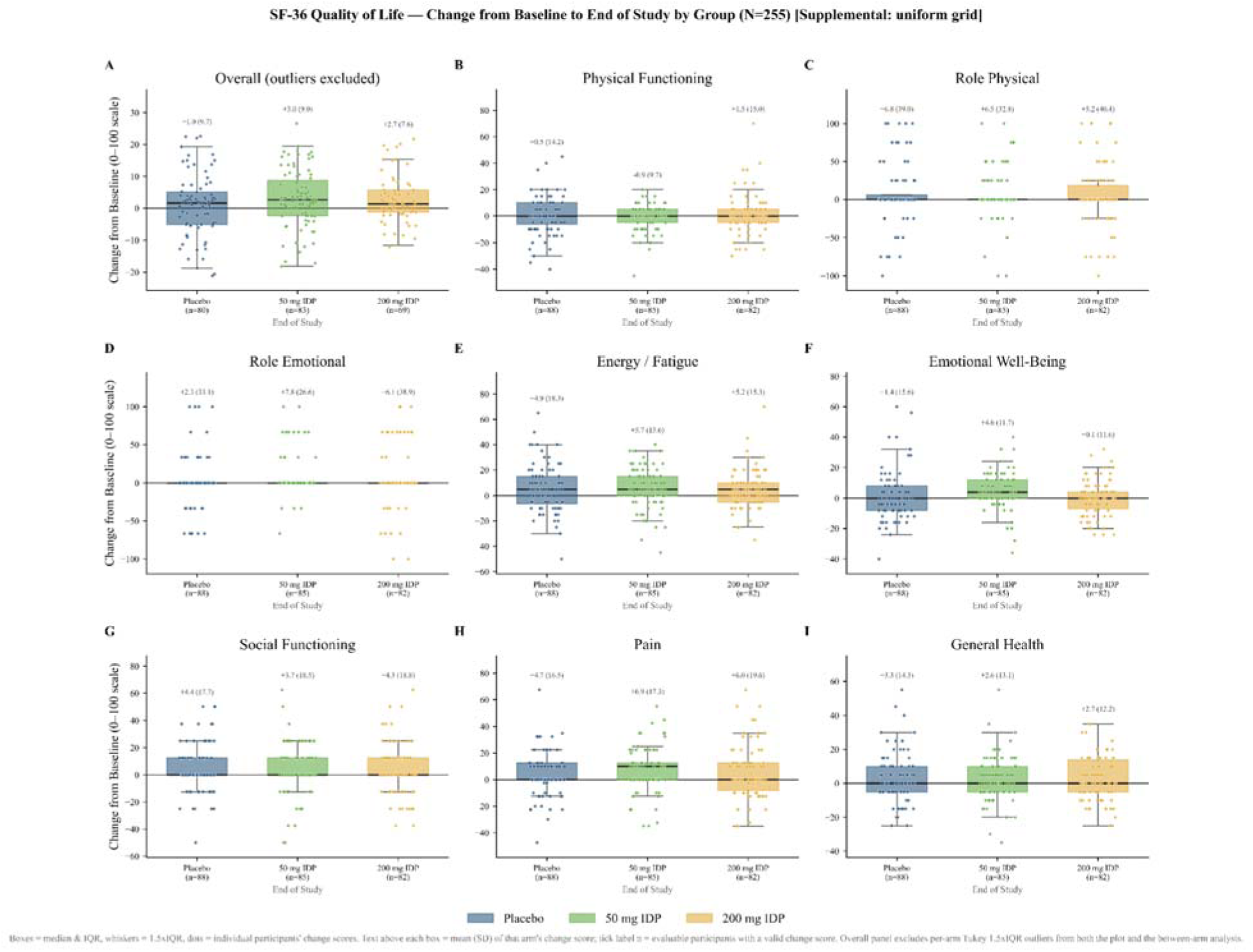
SF-36 Quality of Life Total and Subscore Change From Baseline. Box and whisker plots of change from baseline to end of study in SF-36 A) Total Score, B) Physical Functioning, C) Role Physical, D) Role Emotional, E) Energy, F) Emotional Wellbeing, G) Social Functioning, H) Pain, and I) General Health subscales subscale scores by arm and time. Overlaid scatters represent individual level data. A group-by-time mixed model showed no between-arm differences. Numbers above each box depict mean (SD) of change score by group.

**Supplemental Figure 3.**
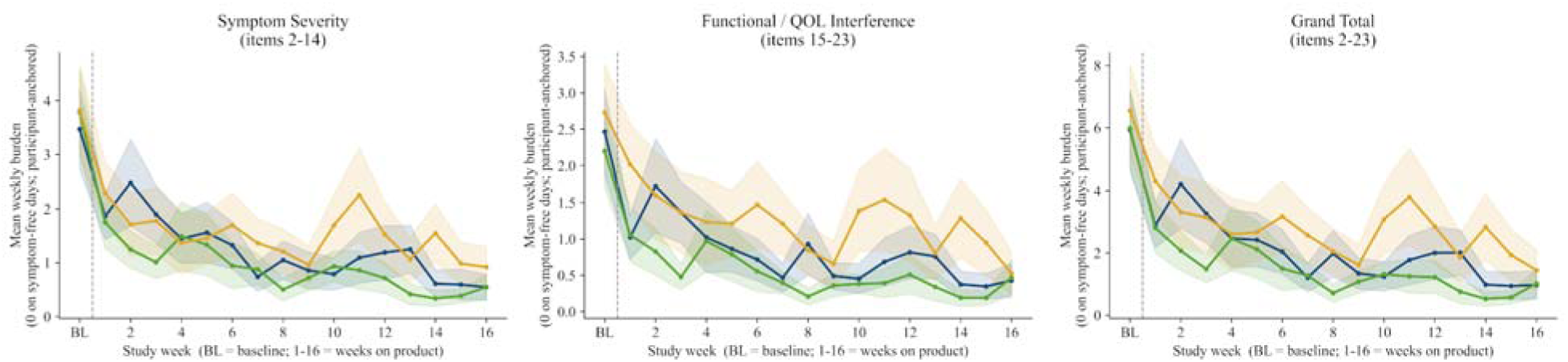
WURSS-24 Symptom Severity Score, QOL Interference Score, and Grand Total Score. Lines indicate mean with shaded SEM, with Placebo in blue, 50 mg IDP® in green and 200 mg IDP® in yellow. Groups did not differ statistically at any time point.

**Supplemental Table 1.**
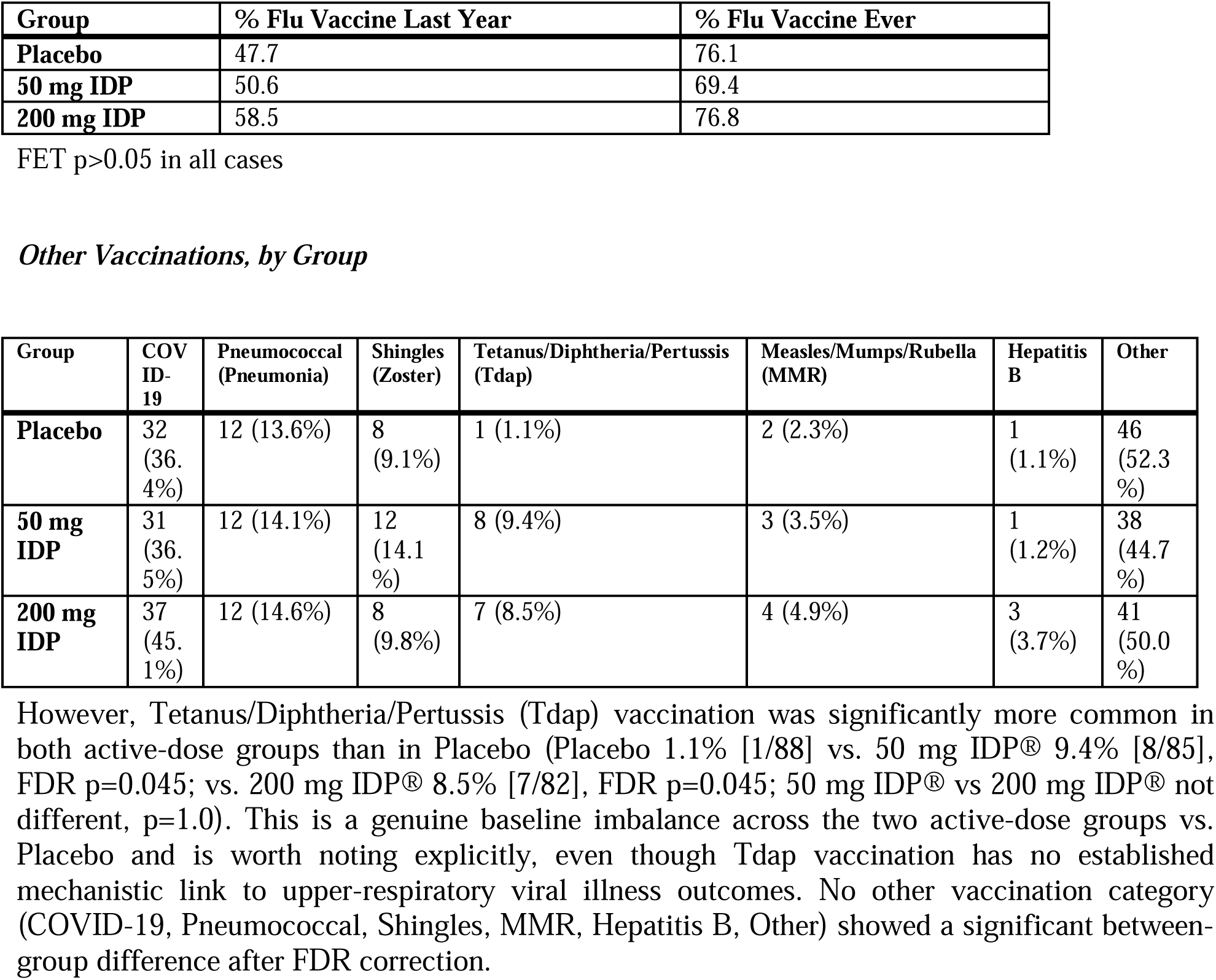
Flu Vaccination Status by Group.

| Group | % Flu Vaccine Last Year | % Flu Vaccine Ever |  |  |  |  |  |
|---|---|---|---|---|---|---|---|
| **Placebo** | 47.7 | 76.1 |  |  |  |  |  |
| **50 mg IDP** | 50.6 | 69.4 |  |  |  |  |  |
| **200 mg IDP** | 58.5 | 76.8 |  |  |  |  |  |
| **Group** | **COVID-19** | **Pneumococcal (Pneumonia)** | **Shingles (Zoster)** | **Tetanus/Diphtheria/Pertussis (Tdap)** | **Measles/Mumps/Rubella (MMR)** | **Hepatitis B** | **Other** |
| **Placebo** | 32 (36.4%) | 12 (13.6%) | 8 (9.1%) | 1 (1.1%) | 2 (2.3%) | 1 (1.1%) | 46 (52.3%) |
| **50 mg IDP** | 31 (36.5%) | 12 (14.1%) | 12 (14.1%) | 8 (9.4%) | 3 (3.5%) | 1 (1.2%) | 38 (44.7%) |
| **200 mg IDP** | 37 (45.1%) | 12 (14.6%) | 8 (9.8%) | 7 (8.5%) | 4 (4.9%) | 3 (3.7%) | 41 (50.0%) |

FET p>0.05 in all cases
*Other Vaccinations, by Group*
However, Tetanus/Diphtheria/Pertussis (Tdap) vaccination was significantly more common in both active-dose groups than in Placebo (Placebo 1.1% [1/88] vs. 50 mg IDP® 9.4% [8/85], FDR p=0.045; vs. 200 mg IDP® 8.5% [7/82], FDR p=0.045; 50 mg IDP® vs 200 mg IDP® not different, p=1.0). This is a genuine baseline imbalance across the two active-dose groups vs. Placebo and is worth noting explicitly, even though Tdap vaccination has no established mechanistic link to upper-respiratory viral illness outcomes. No other vaccination category (COVID-19, Pneumococcal, Shingles, MMR, Hepatitis B, Other) showed a significant between-group difference after FDR correction.

